# Comparative Analysis of COVID-19 Transmission Patterns in Three Chinese Regions *vs*. South Korea, Italy and Iran

**DOI:** 10.1101/2020.04.09.20053223

**Authors:** Junyu He, Guangwei Chen, Yutong Jiang, Runjie Jin, Mingjun He, Ashton Shortridge, Jiaping Wu, George Christakos

## Abstract

**Background:** The outbreak of Coronavirus 2019 (COVID-19) began in January 2020 in the city of Wuhan (Hubei province, China). It took about 2 months for China to get this infectious disease under control in its epicenter at Wuhan. Since February 2020, COVID-19 has been spreading around the world, becoming widespread in a number of countries. The timing and nature of government actions in response to the pandemic has varied from country to country, and their role in affecting the spread of the disease has been debated.

**Method:** The present study proposed a modified susceptible-exposed-infected-removed model (SEIR) model to perform a comparative analysis of the temporal progress of disease spread in six regions worldwide: three Chinese regions (Zhejiang, Guangdong and Xinjiang) *vs*. three countries (South Korea, Italy and Iran). For each region we developed detailed timelines of reported infections and outcomes, along with government-implemented measures to enforce social distancing. Simulations of the imposition of strong social distancing measures were used to evaluate the impact that these measures might have had on the duration and severity of COVID-19 outbreaks in the three countries.

**Results:** The main results of this study are as follows: (*a*) an empirical COVID-19 growth law provides an excellent fit to the disease data in all study regions and potentially could be of more general validity; (*b*) significant differences exist in the spread characteristics of the disease among the three regions of China and between the three regions of China and the three countries; (*c*) under the control measures implemented in the Chinese regions (including the immediate quarantine of infected patients and their close contacts, and considerable restrictions on social contacts), the transmission rate of COVID-19 followed a modified normal distribution function, and it reached its peak after 1 to 2 days and then was reduced to zero 11, 11 and 18 days after a 1^st^-Level Response to Major Public Health Emergency was declared in Zhejiang, Guangdong and Xinjiang, respectively; moreover, the epidemic control times in Zhejiang, Guangdong and Xinjiang showed that the epidemic reached an “inflection point” after 9, 12 and 17 days, respectively, after a 1^st^-Level Response was issued; (*d*) an empirical COVID-19 law provided an excellent fit to the disease data in the six study regions, and the law can be potentially of more general validity; and (*e*) the curves of infected cases in South Korea, Italy and Iran would had been significantly flattened and shrunken at a relatively earlier stage of the epidemic if similar preventive measures as in the Chinese regions had been also taken in the above three countries on February 25^th^, February 25^th^ and March 8^th^, respectively: the simulated maximum number of infected individuals in South Korea, Italy and Iran would had been 4480 cases (March 9^th^, 2020), 2335 cases (March 10^th^) and 6969 cases (March 20^th^), instead of the actual (reported) numbers of 7212 cases (March 9^th^), 8514 cases (March 10^th^, 2020) and 11466 cases (March 20^th^), respectively; moreover, up to March 29^th^, the simulated reduction in the accumulated number of infected cases would be 1585 for South Korea, 93490 for Italy and 23213 for Iran, respectively, accounting for 16.41% (South Korea), 95.70% (Italy) and 60.59% (Iran) of the accumulated number of actual reported infected cases.

**Conclusions:** The implemented measures in China were very effective for controlling the spread of COVID-19. These measures should be taken as early as possible, including the early identification of all infection sources and eliminating transmission pathways. Subsequently, the number of infected cases can be controlled at a low level, and existing medical resources could be sufficient for maintaining higher cure rates and lower mortality rate compared to the current situations in these countries. The proposed model can account for these prevention and control measures by properly adjusting its parameters, it computes the corresponding variations in disease transmission rate during the outbreak period, and it can provide valuable information for public health decision-making purposes.

## 1. Introduction

The Coronavirus 2019 (COVID-19) is an RNA virus that has a 79.5% nucleic acid similarity with the Severe Acute Respiratory Syndrome (SARS) coronavirus and a 96% nucleic acid similarity with the bat coronavirus [1]. At present, it is believed that the main transmission routes of COVID-19 are respiratory droplets and direct contact [2]. In addition, it has the potential to spread through aerosols and the fecal-oral route [3]. In particular, COVID-19 has been found to be floating in a closed carriage for 30 minutes over distances of up to 4.5 meters during long-distance passenger transport. The S protein of COVID-19 fuses to the full-length protein of human ACE2 and invades the human body, causing illness [4]. Fever and cough are COVID-19’s two main clinical symptoms [2]. Moreover, the patient may also experience fatigue, muscle soreness, phlegm, headache, hemoptysis, diarrhea and other symptoms [5]. The pathological features of COVID-19 are very similar to those seen in patients with SARS and Middle East respiratory syndrome (MERS) [6]. The pulmonary fibrosis is not too severe, but the inflammation is severe and there is a lot of mucus. Due to the incubation period of COVID-19, if effective measures are not taken to control the source of infection and cut off the transmission route, it will lead to the rapid spread of the disease. Since the COVID-19 outbreak in Wuhan city in January 2020, nearly 67,000 cases had been confirmed in city residents by March 30, nearly 63,000 had been cured, and more than 3,000 had died [7]. To date, more than 82,000 people have been confirmed as COVID-19 infected in China, nearly 76,000 have been cured, and more than 3,300 have died nation-wide [7].

In response to the COVID-19 outbreak, the Chinese authorities acted quickly by taking strict prevention and control measures that focused on Wuhan city [7-9], and by providing the necessary support in terms of medical teams, instruments, other supplies and funds. A detailed outline and timetable of the Chinese efforts is given in Fig S1 in the Supporting Material section. The motto “one province (in the Country) assists one city in Hubei province” was implemented in an effort to support the medical team in key epidemic areas. After nearly two months of fighting the disease nation-wide, by March 18^th^, 2020 the newly confirmed COVID-19 patients had dropped to zero in mainland China. It should be also noticed that some infected individuals traveled to China from abroad. For example, a COVID-19 case in Ningxia was reported from abroad on February 26, 2020. By March 29^th^, a total of 723 cases were confirmed.

At the time of writing, over 1.5 million COVID-19 cases have been recorded in more than 100 countries worldwide. By March 29^th^, over 0.66 million infected cases had been confirmed worldwide, among whom 0.14 million people have been cured and 30 thousand people have died. Interestingly, the measures taken in China have reduced the disease’s spread. For example, studies have shown that the blockade of Wuhan city reduced international transmission by 80% by mid-February, 2020 [11]. Next, certain characteristics of the COVID-19 spread in six selected regions worldwide are discussed, comparisons are made between them, and the effects of prevention and control measures are investigated. The results could valuably inform future strategies of epidemic prevention and control.

## 2. Materials and methods

### 2.1 Study regions and data collection

In this work, the COVID-19 spread patterns in three regions of China were compared with the corresponding disease patterns in three selected countries, as follows:

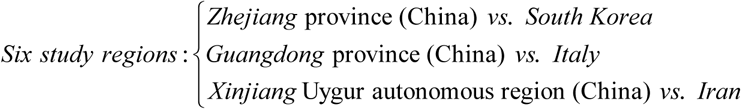

These comparison choices were made on the basis of the epidemic features observed in the corresponding regions, as well as on socio-economic and environmental criteria. Around February 20^th^, when South Korea, Italy and Iran showed signs of potential COVID-19 outbreaks, Zhejiang and Guangdong already have had the highest numbers of COVID-19 cases in China (outside of Hubei province). Italy, South Korea and Iran were affected relatively early in the pandemic and developed a large number of confirmed COVID-19 cases. Also, as is discussed in [12-15], there exist certain similarities in the social, economic or environmental conditions between the pairs “Zhejiang-South Korea,” “Guangdong-Italy,” and “Xinjiang-Iran”. Zhejiang and Guangdong are among the most internationalized regions in China. Zhejiang and South Korea have almost the same land area and population, and their climate and GDP are relatively close. Among all provinces and autonomous regions in China, Guangdong is the one that could probably best match Italy, by the overall consideration of social, economic and environmental conditions. Xinjiang and Iran have social and culture similarities. Detailed descriptions of the six study regions can be found in the Supporting Materials section (Text S1). The numbers of infected, cured and dead individuals were collected from the Health Commissions of the three regions in China [16-18] and some websites documented the corresponding numbers for Italy, South Korea and Iran [19-21].

### 2.2 Methods

The methodology of the present study consisted of two parts: (*a*) the disease spread was represented mathematically by an epidemic model, and (*b*) the effects of the measures taken by the various regions to control the disease were compared in terms of the model parameters.

Many models linking the evolution of susceptible, infected and removed cases have been used in the study of infectious disease distributions [22-26]. A modified susceptible-exposed-infected-removed model (SEIR) was used in this study to simulate the COVID-19 spread in the six study regions. The basic outline of the SEIR model is shown in Fig S2, and the corresponding equations are as follows:

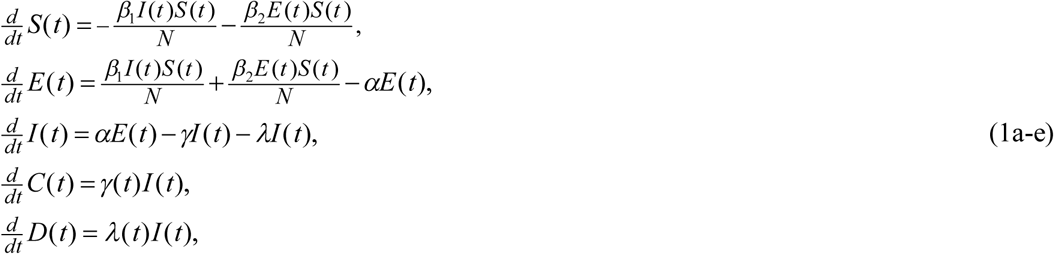

where *N* denotes the population sizes of the regions of interest, *S*(*t*), *E*(*t*), *I*(*t*), *C*(*t*) and *D*(*t*) represent, respectively, the number of susceptible, exposed (may or may not become infected), infected, cured and dead individuals at time *t*. The exposed and infected individuals constitute the total number of affected individuals *A*(*t*) = *E*(*t*) + *I* (*t*). The cured and dead individuals constitute the total number of removed individuals *R*(*t*) = *C*(*t*) + *D*(*t*). The time-varying parameters *β*_1_(*t*) and *β*_2_(*t*) denote the rate of COVID-19 transmission when an individual comes in contact with infected and exposed individuals, respectively, the constant α represents the probability that the exposed individuals become infected, and the time-varying parameters *γ* (*t*) and *λ*(*t*) denote the COVID-19 cure rate and death rate, respectively. The SEIR model was developed specifically for this study because the COVID-19 incubation period is about 7 days, and early in this period no symptoms are detected, which means the infected but asymptomatic people will unknowingly infect others before they develop any symptoms.

According to the proposed SEIR model, the infection contact rate 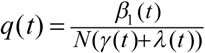 expresses the fraction of population that comes into contact with an infected individual during the infection period, and an expanding COVID-19 epidemic may be expected to occur at any time *t* in a region when the following inequality holds:

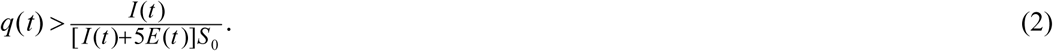

Alternatively, the *R*_0_(*t*) = *q*(*t*)*S*_0_ represents the number of secondary infections at any time *t* in the population caused by an initial primary infection. If one person has COVID-19, *R*_0_(*t*) determines how many infections on average that person may cause. Then, the epidemic would be considered under control at the time *t*_*ECT*_ (epidemic control time, ECT) after which the *R*_0_(*t*) is consistently smaller than *L*_0_(*t*), i.e., the inequality

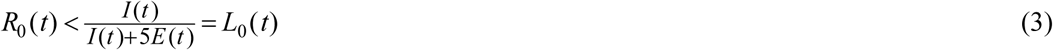

holds for all *t* ≥ *t*_*ECD*_.

The SEIR model parameters above are very important because they determine (to varying extents and within different contexts) the epidemic spread and its severity. Therefore, identifying good estimates is critical for realistic simulation of infectious disease. In this work, we classified the SEIR parameters into two groups: *Preventative* and *post-infection*. The infection transmission rate *β*_1_(*t*), the exposure transmission rate *β*_2_(*t*) and the infection probability *α* of exposed individuals are *preventive* parameters and their values depend on the measures taken by responsible individuals and authorities prior to disease exposure (e.g., hygiene, disinfection, protective equipment, social distancing, testing, isolation, etc.). The cure rate *γ* (*t*) and the death rate *λ*(*t*) are *post-infection* parameters that depend on a range of disease and social factors (e.g., lethality of the pathogen, pre-existing health condition of the infected individuals, the timeliness and quality of health care available etc.)

One of the objectives of this work is to investigate how the COVID-19 epidemic can be better understood and how its control might be improved by adjusting the corresponding SEIR model parameters according to the disease prevention and control measures listed above. Otherwise said, to prevent or control an epidemic means to control the model parameters such as *β*_1_(*t*), *β*_2_(*t*) and *α*. In this work, then, the following modeling and computational choices were made associated with the SEIR of Eqs (1a-e).

i. The sum of the number of infected, cured and dead individuals at time *t* is equal to the accumulated number of confirmed infected individuals at the same time. The populations *N* of the regions and countries of interest were assumed to be large enough that they can be assumed to remain constant during the epidemic. Some interesting inequalities hold, like *S*(*t*) ≤ *S*_0_, i.e., the susceptible population *S*(*t*) at any time is smaller than the initial number of susceptible cases (*S*0).
ii. In the SEIR equations, the exposure rate includes two parts: one part is proportional to the contacts between susceptible and infected individuals (assessed by *β*_1_), whereas the other part is proportional to the contacts between susceptible and exposed individuals (assessed by *β* _2_). Since individuals that are confirmed as infected are immediately sent to hospitals for isolation and medical treatment in the three regions of China, it is assumed that the infection transmission rate was always smaller than the exposure transmission rate, i.e., *β*_1_ < *β*_2_. Specifically, following the current COVID- 19 literature [27], it is assumed that *β*_2_ = 5*β*_1_, and that the probability that an exposed individual becomes infected is 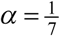. The cured and dead individuals are removed at the rate *γ* (*t*) ± *λ* (*t*). In the regions of interest, the values of *γ* and *λ* were zero at the beginning (*t* = 0), i.e., *γ*_0_ = *λ*_0_ = 0.
iii. Given that the COVID-19 disease spread has been controlled in the three regions of China, only data from the first 28-days were taken into account in the present study. For South Korea, Italy and Iran, complete datasets covering the entire countries were considered, including areas in which epidemic spread is still underway.
iv. Transmission, cure, and mortality rates are unknown and dynamic. We estimate these parameters for each region using particle swarm optimization (PSO), a nonlinear computational fitting procedure [28,29]. A 6-days long moving window was introduced for computational model parameter fitting (i.e., the infection transmission rate, *β*_1_, the cure rate, *γ*, and the mortality rate, *λ*). The computational modeling procedure included the steps listed in Table 1. The actual (empirical) values of the three rates varied during the 6-days window period, but in Table 1 the values of the three PSO- fitted parameters are assumed constant, corresponding to the minimum residuals (actual *vs*. fitted numbers of infected cases, cured cases and dead cases) during the same period. The *β*_1_, *γ* and *λ* values at the regions of interest were compared by considering the effective measures taken by the various regions to control the disease. In addition, considering that the increasing number of infected individuals in China has become very small, the COVID-19 transmission pattern (law) adopted by the Chinese authorities was investigated by fitting the transmission rate curve to empirical mathematical models.

## 3 Results

### 3.1 Comparative analysis of disease spread *vs*. preventive measures in the six regions

Fig 1 shows the accumulated and cured cases in the six regions considered with the major disease control and prevention events that took place at each of the six regions. Moreover, below we provide more details about the situation:

**Table 1:**
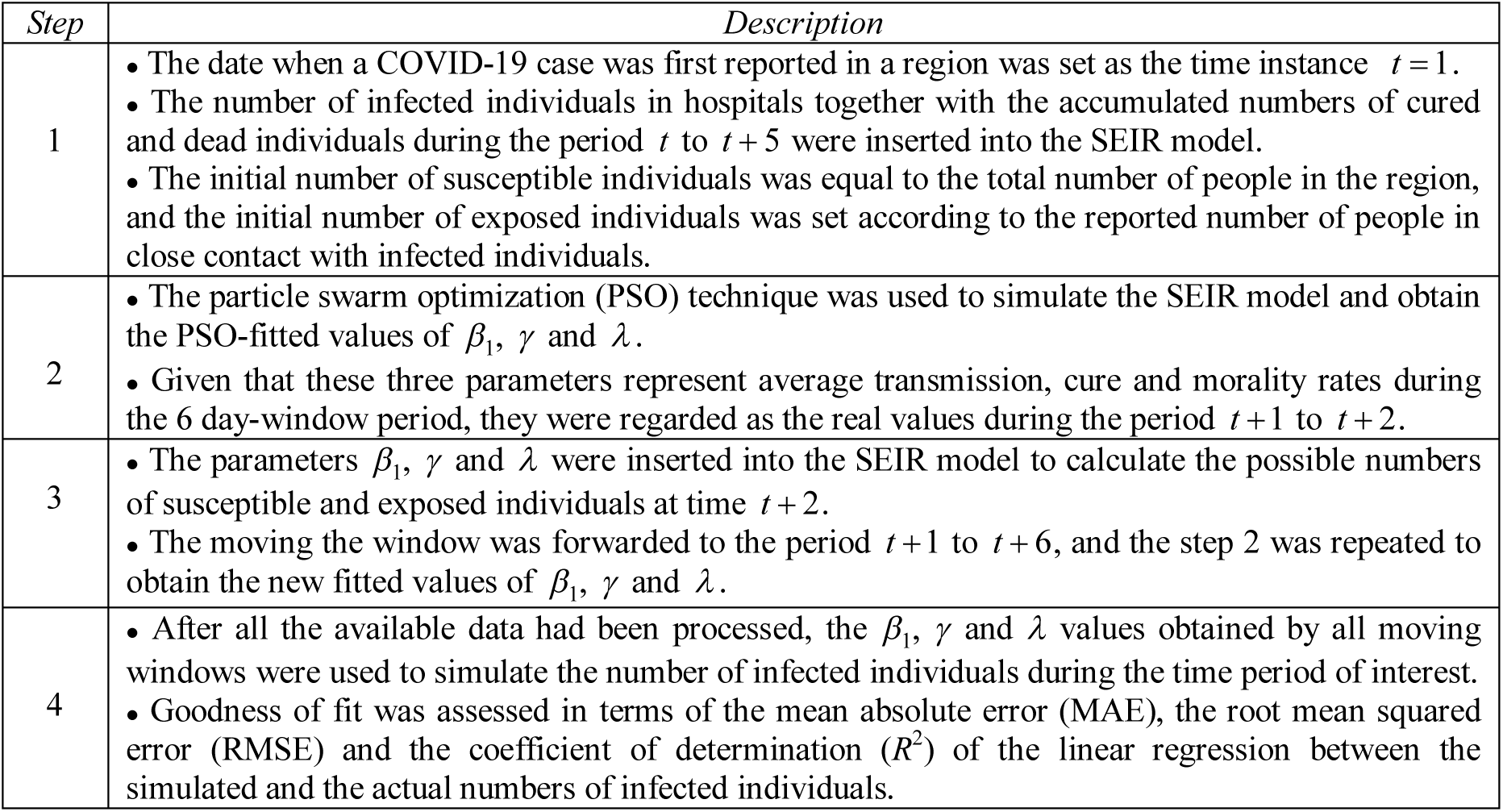
The SEIR computational modeling procedure.

**Figure 1:**
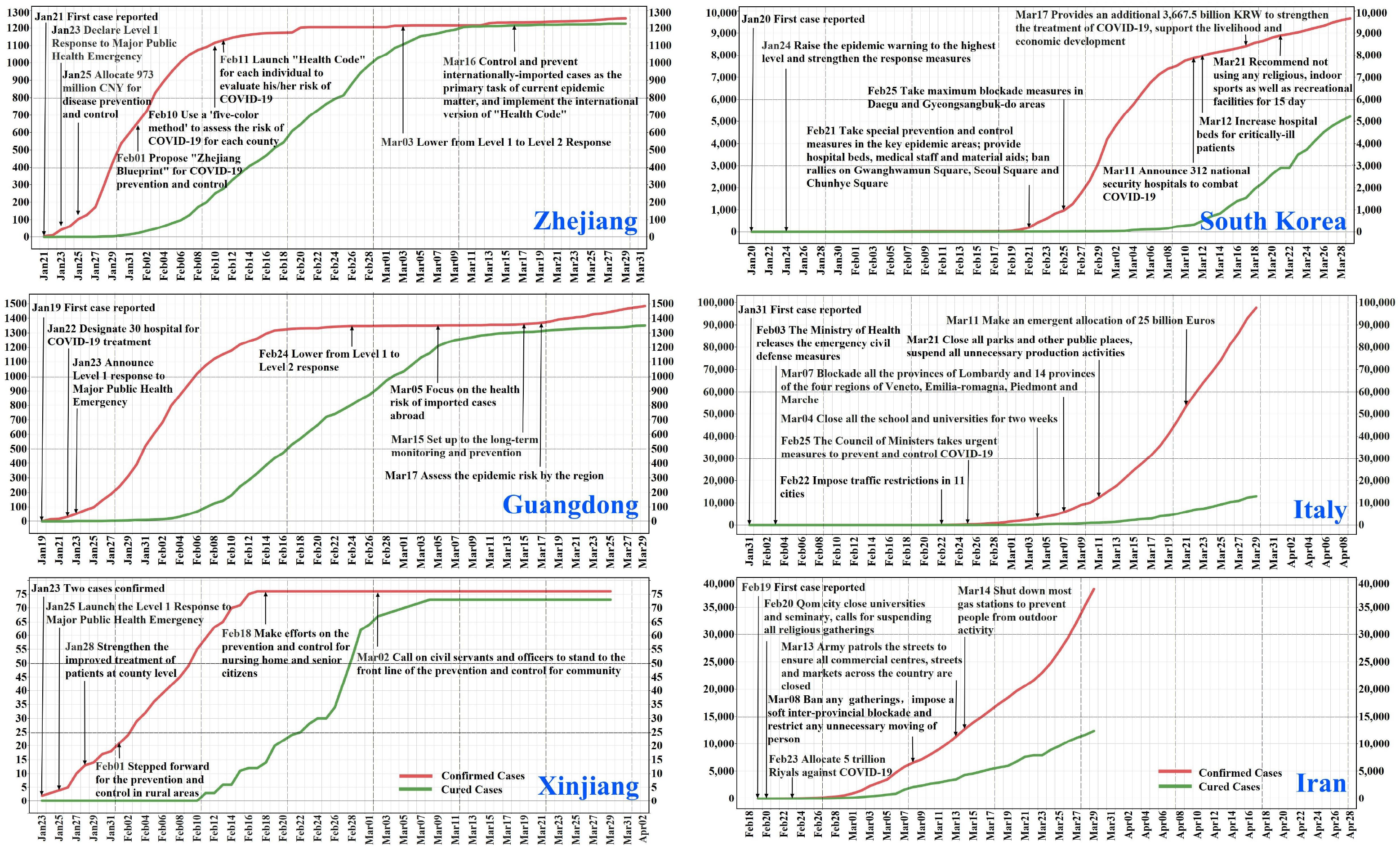
Temporal evolution of accumulated and cured cases of COVID-19 diseases in Zhejiang, Guangdong and Xinjiang of China, South Korea, Italy and Iran. Major events about the disease control and prevention are labeled at the corresponding date.

#### Zhejiang

Zhejiang announced its 1^st^-level Response to Major Public Health Emergency on January 23^rd^, 2020, two days after its first COVID-19 case was confirmed. During the period January 28^th^-30^th^, 364 patients were confirmed, which was 2.1 times higher than the number of confirmed cases during the period January 21^th^-27^th^. On the other hand, the COVID-19 incubation period delayed the outbreak. Since February 11^th^, a “Health Code” was launched to evaluate the health risk of COVID-19. Within three weeks since the first reported case in the region, a plateau of about 1200 cumulative cases was reached and only one death was reported on February 19^th^. Around March 10^th^, nearly all cases were cured with an average stay of 18.17 days in hospital. Due to more and more cases confirmed from international travelers, on March 16^th^, the control and prevention of internationally imported cases became the primary task, and an international version of “Health Code” was implemented to assess the health risk of individuals entering Zhejiang.

#### South Korea

Even before the first COVID-19 case was confirmed in South Korea, the public health department started developing a COVID-19 testing method on Jan 13^th^, 2020 [19]. On January 30^th^, a new real-time COVID-19 detection technique was developed and validated that has an outstanding detection speed and feasibility [19]. During the period from January 20^th^ to Feb 18^th^, South Korea had a relatively low number of confirmed COVID-19 cases (Fig 1). However, because of high density gathering involving infected people, the number of confirmed cases increased a lot after that date. The numbers confirmed cases suddenly exploded, from 104 cases on February 20^th^ to 7979 cases on March 12^th^, which further increased to 9661 cases on March 29^th^. In particular, in Daegu City, the number of infected cases was raised to 710 on February 26^th^, which was more than half of the total number of cases in South Korea at that time. On February 25^th^, the South Korean government imposed the strictest blockades in Daegu city and North Gyeongsang province. After that, disease transmission has been gradually controlled. On March 10^th^, the Central Disaster and Safety Countermeasure Headquarters conducted an on-site inspection of nursing hospitals seeking to block regional small group transmission [19]. Note that until March 31^th^, 2020, there were 339 hospitals treating the COVID-19 patients in South Korea [30].

#### Guangdong

On January 22^nd^, 2020, Guangdong set up 30 hospitals for exclusively treating COVID-19 infected individuals. On January 23^rd^, the local government announced the 1^st^-level Response to Major Public Health Emergency. According to Fig 1, the number of confirmed newly infected individuals on a daily basis was very large at the beginning of the disease outbreak, but gradually became smaller, which means that the disease control measures taken in Guangdong were indeed effective. By February 15^th^, 2020, a total of 1316 individuals were confirmed as COVID-19 infected. The local government lowered the level response to the public health emergency event on February 24^th^. Since February 28^th^, the local authorities focused on curing two classes of patients (severe and critically ill individuals), and on reducing the mortality rate by dividing the hospitals into two groups, according to the above two patient classes. Since March 5^th^, the main focus of the local authorities has been on imported cases from abroad.

#### Italy

In Italy (Fig 1), the first COVID-19 infected individual was confirmed on January 31^st^, 2020, i.e., 10 days later than in Guangdong. In the following two days, the Italian government declared a state of emergency in the country. On Feb 22^nd^, the COVID-19 began to be prevalent; the Italian government immediately restricted the traffic between 11 cities. Production activities and public gatherings were paused and schools were closed in these cities. To interrupt the transmission path, on March 7^th^ the government blockaded all the provinces in Lombardy and the 14 provinces of the four regions of Veneto, Emilia-Romagna, Piedmont and Marche.

#### Xinjiang

]The total number of confirmed COVID-19 cases during the study period was low (76), but with a relatively high mortality of 3.95% (Fig 1). On January 25^th^, the 1^st^-level Response to Major Public Health Emergency was issued in the region. The disease control and prevention measures were shifted from the city to the county level and then to the rural areas.

#### Iran

The first COVID-19 case was reported on February 19^th^, 2020 (Fig 1). One day later, the local government of Qom City (Iran) closed the universities and the schools. On March 8^th^, Iran issued a fatwa to ban any religious gathering, impose a soft inter-provincial blockade, and restrict any unnecessary movement of people. Starting March 13^th^, 2020, the Iranian army was deployed to ensure that the commercial centers, streets and markets were kept closed.

### 3.2 Comparative analysis of the case increment trends in the six regions

Fig 2 is a plot of the daily numbers of newly infected cases as a function of the accumulated numbers of infected cases in the three Chinese regions and the three countries. Interestingly, the dots representing the Zhejiang, Guangdong, Xinjiang and South Korea cases showed that after they reached their peaks, the cases exhibited decreasing trends as the accumulated number of infected cases increased further. On the other hand, the dots representing Italy and Iran continue to exhibit increasing trends as functions of the accumulated number of infected cases. The initial increasing trends are similar for all regions considered, but then significant differences occurred. The Xinjiang curve was the first to show a downward decline (implying a rather early epidemic control), followed by the Zhejiang and Guangdong curves at later times (these two curves are remarkably similar). The South Korean curve initially followed the linear increase of the other curves, it then briefly experienced a temporary downwards slope, but it soon started increasing again; finally, the South Korean declined (indicating that control of the epidemic developed at a later time). The Italian and Iranian curves continued increasing, emphasizing ongoing worsening epidemic situations in these two countries during the study period.

**Figure 2:**
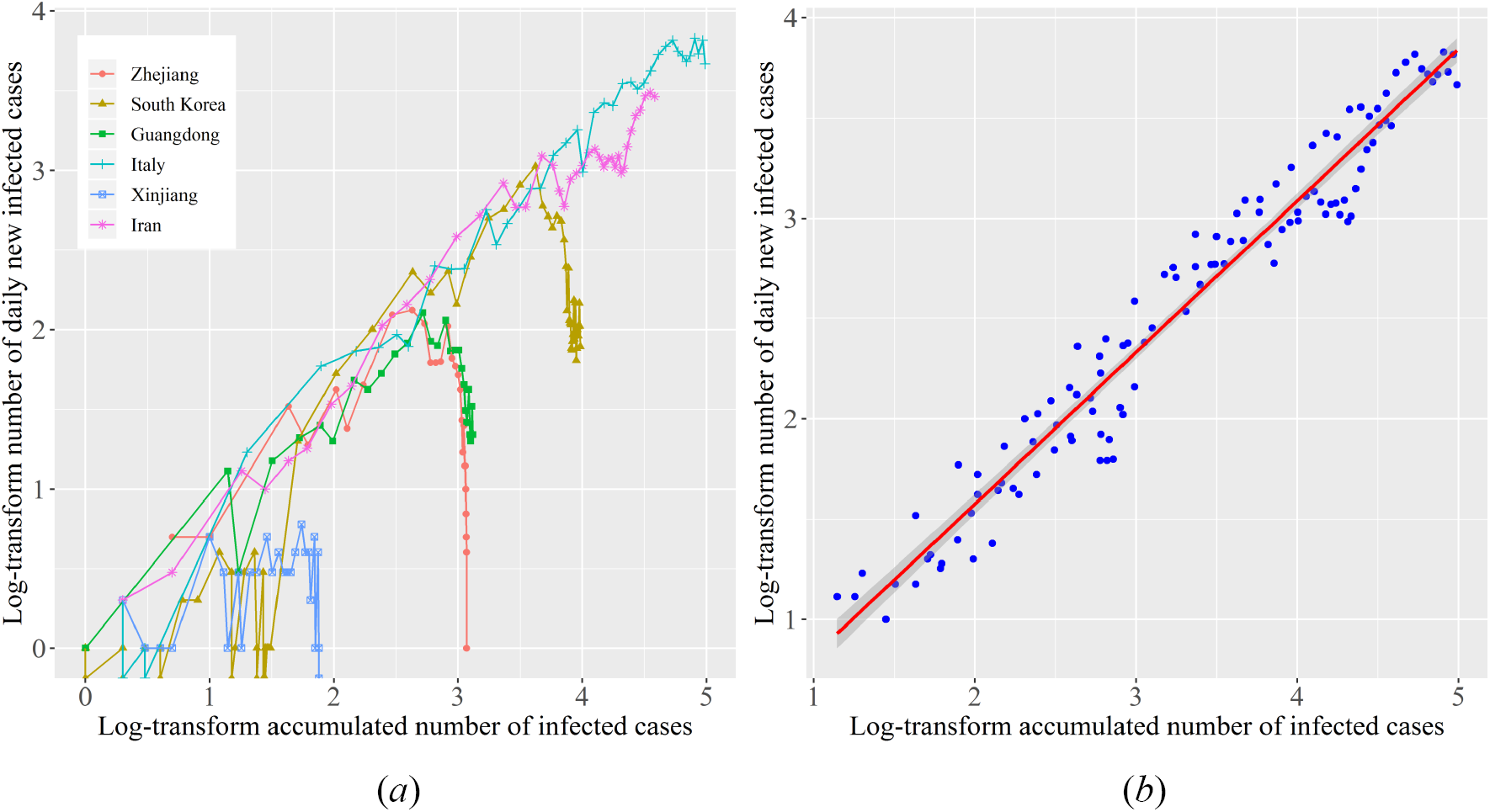
Relationship between the numbers of daily new infected cases *vs*. the accumulated numbers of infected cases. (*a*) Scatter-plots of this relationship for the six regions; (*b*) linear fitting of the scatter-plots with 95% confidence interval shown as a shaded area (excluding the scatter points showing declining trends or with the number of daily new infected cases less than 10).

It was found that during the critical growth period of the epidemic (Fig 2b), the COVID-19 variation in all six regions obeyed the log- linear relationship

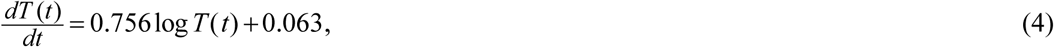

where *T* (*t*) = *I* (*t*) + *R*(*t*). Eq (4) reflects the transmission speed of the COVID-19. Given that Eq (4) fits the data with such a high accuracy, *R*^2^ = 0.96 and *p* < 2.2 ×10^−16^ ≈ 0, it can be viewed as COVID- 19 growth law linking the log-number of daily new infected cases, 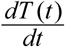, and the log-accumulated number of the infected cases *T* (*t*). That is, this is a general law that potentially applies to COVID-19 epidemics in other regions of the world, and as such it could be a useful tool in future COVID-19 investigations.

#### 3.3 Comparison of the COVID-19 transmission patterns in the six regions

SEIR model parameters were estimated by introducing the number of infected, cured, and dead individuals during the first 28 days of the COVID-19 outbreak in each of the three Chinese regions (Zhejiang, Guangdong and Xinjiang). Variations of the empirical transmission, cure and mortality (death) rates in the three regions at various times are denoted by dots in the upper left parts of Figs 3, 4 and 5, respectively, (the corresponding fitted lines are also shown). Note that in these figures four kinds of plots are shown:

a. actual or empirical (transmission, cure and mortality) rates and the lines fitted to them (for all six regions),
b. simulated transmission rates that should be assumed in the three countries (South Korea, Italy and Iran) if the control and prevention measures of the corresponding Chinese regions had been used,
c. empirical case numbers (infected cases, accumulated cured cases and accumulated death cases) together with the case number plots produced by the SEIR model (for all six regions), and
d. simulated disease case number plots that would be obtained in the three countries (South Korea, Italy and Iran) if the simulated transmission rates corresponding to the control measures of the Chinese regions had been used.

**Figure 3:**
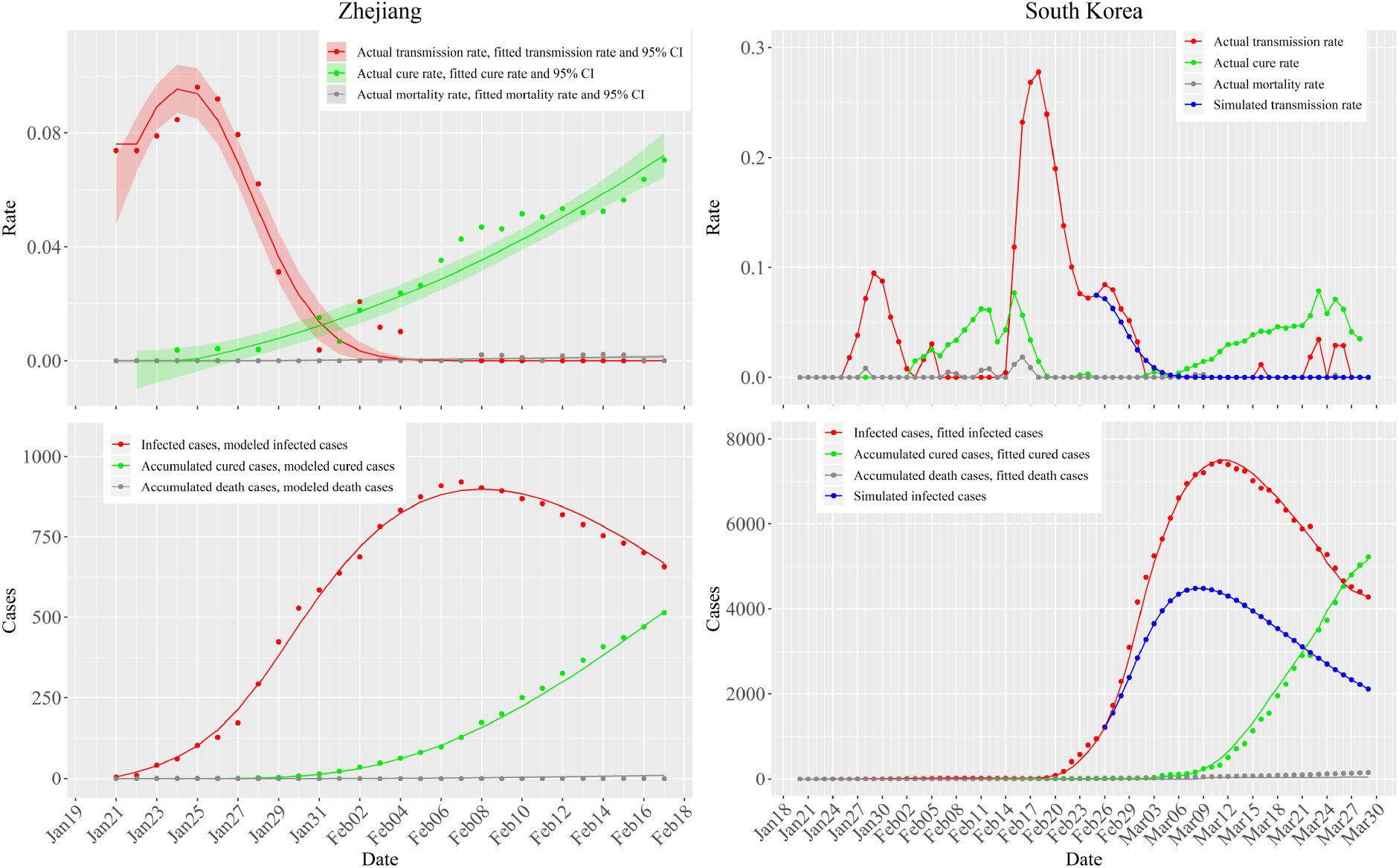
SEIR modeling results in Zhejiang (left column) and in South Korea (right column). The upper figures show the empirical values of transmission rate (red dots), cure rate (green dots) and mortality rate (gray dots) together with the corresponding fitted lines and the 95% confidence interval (shadows); the bottom figures show the reported number of infected cases (red dots), accumulated numbers of cured cases (green dots) and dead cases (gray dots) together with the corresponding SEIR-produced values (lines); graphs for South Korea also presents the simulated rates (upper, blue dots and lines) and number of infected cases (bottom, blue dots and lines) based on a simulation of Zhejiang-style control measures taken since February 25^th^, 2020

**Figure 4:**
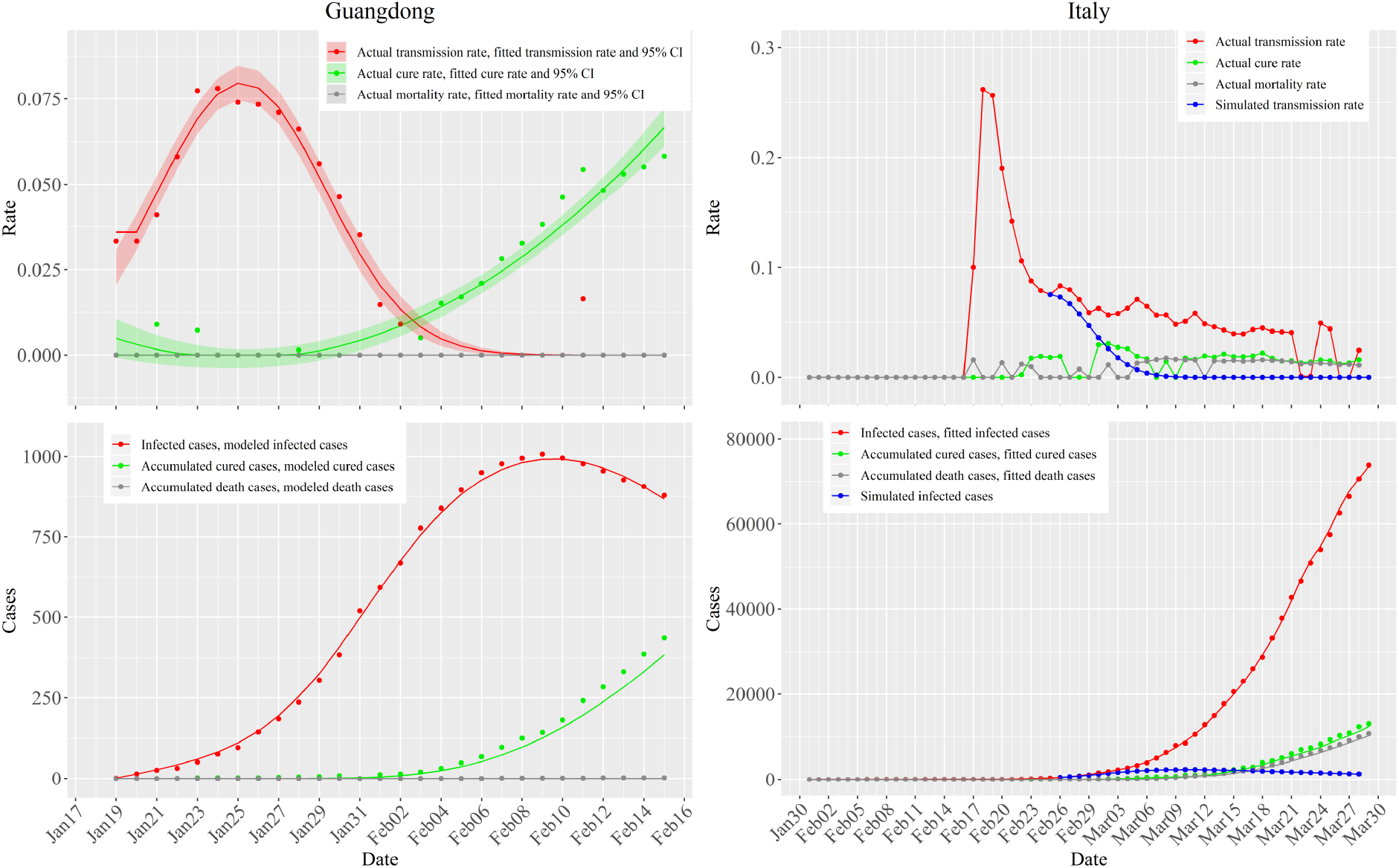
SEIR modeling results in Guangdong (left column) and in Italy (right column). The upper figures show the empirical values of transmission rate (red dots), cure rate (green dots) and mortality rate (gray dots) together with the corresponding fitted lines and the 95% confidence interval (shadows); the bottom figures show the reported number of infected cases (red dots), accumulated numbers of cured cases (green dots) and dead cases (gray dots) together with the corresponding SEIR-produced values (lines); graphs for Italy also presents the simulated rates (upper, blue dots and lines) and number of infected cases (bottom, blue dots and lines) based on a simulation of Guangdong-style control measures taken since February 25^th^, 2020

**Figure 5:**
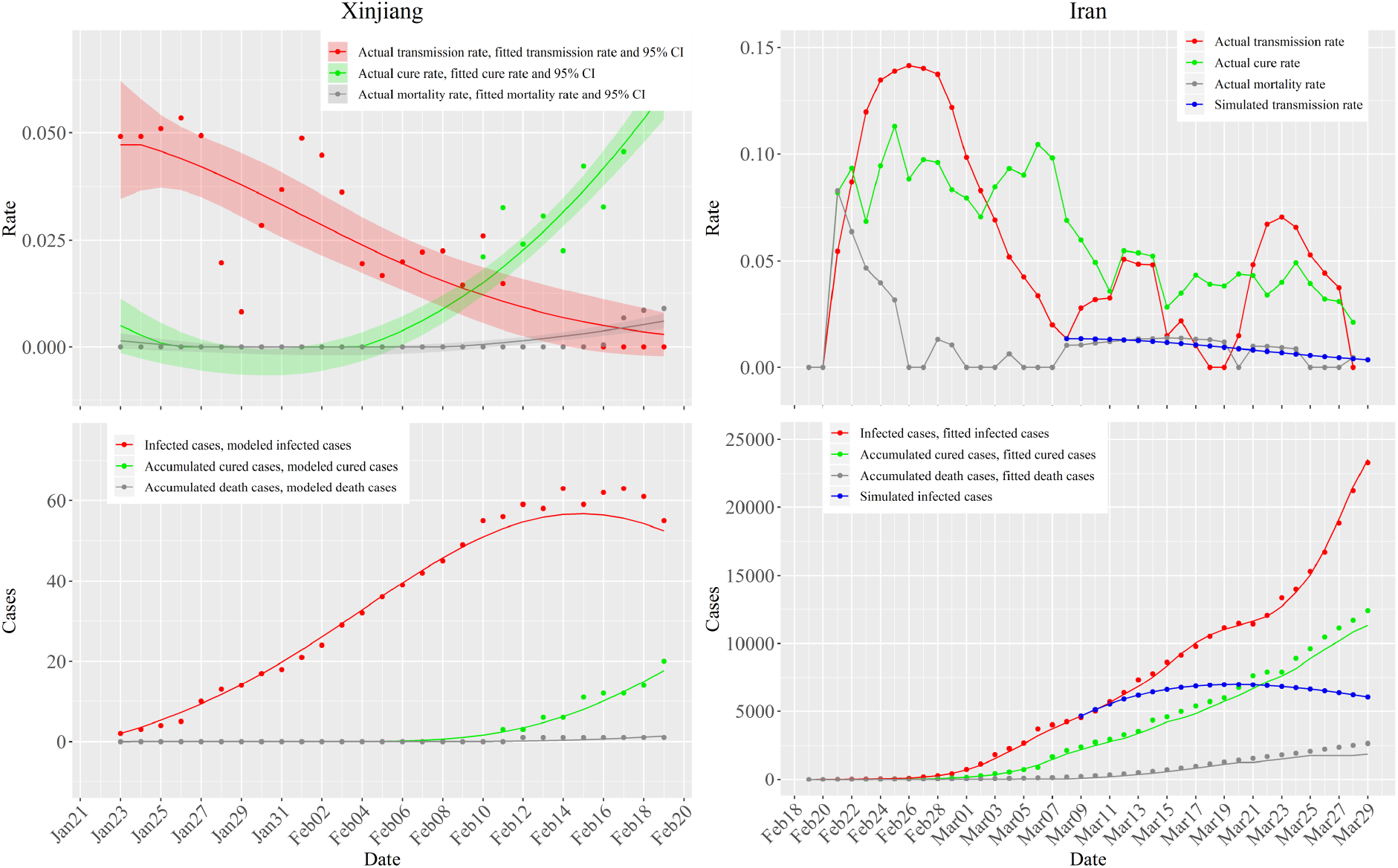
SEIR modeling results in Xinjiang (left column) and in Iran (right column). The upper figures show the empirical values of transmission rate (red dots), cure rate (green dots) and mortality rate (gray dots) together with the corresponding fitted lines and the 95% confidence interval (shadows); the bottom figures show the reported number of infected cases (red dots), accumulated numbers of cured cases (green dots) and dead cases (gray dots) together with the corresponding SEIR-produced values (lines); graphs for Iran also presents the simulated rates (upper, blue dots and lines) and number of infected cases (bottom, blue dots and lines) based on a simulation of Xinjiang-style control measures taken since March 8^th^, 2020

The empirical cure rates at the three regions exhibited increasing trends, whereas the mortality rates varied: 0.08% in Zhejiang, 0.59% in Guangdong to 3.95% in Xinjiang. Regarding the empirical transmission rates, Zhejiang and Guangdong showed similar increasing-decreasing trends. The empirical transmission rates at Xinjiang showed an increasing-decreasing trend but with certain noticeable fluctuations. The transmission rate peaks were reached within 1 day in both Guangdong and Xinjiang, and 2 days in Zhejiang after the 1^st^-Level Response to Major Public Health Emergency was launched at, respectively, Zhejiang (January 23^th^ 2020), Guangdong (January 23^th^) and Xinjiang (January 25^th^). The highest transmission rates were 0.096 (Zhejiang), 0.075 (Guangdong), and 0.053 (Xinjiang). The empirical transmission rate dropped to zero after 11 days (Zhejiang), 11 days (Guangdong) and 18 days (Xinjiang) since the 1^st^-Level responses were launched in these regions.

Given the shapes of the three rate curves for Zhejiang, Guangdong and Xinjiang, the modified normal distribution function (Text S2 in the Supporting Material section) was fitted to the empirical transmission rates, whereas the quadratic function was fitted to the empirical cure and mortality rates (more detailed information can be found in the Supporting Material section). The three fitted curves (denoted by lines) together with their 95% confidence intervals (denoted by shadow areas) depicting the variation pattern of the empirical transmission, cure and mortality rates are shown at the top left part of Figs 3, 4 and 5, respectively. Using the fitted rate curves above, the case numbers of these three COVID-19 groups of individuals (infected, cured and dead) generated by the SEIR model in Zhejiang, Guangdong and Xinjiang are denoted by continuous lines at the bottom-left of Figs 3 to 5, whereas the actual case numbers of the three groups of individuals are denoted by dots (the *R*^2^ values of the number of infected individuals vary from 0.980 to 0.999). More detailed goodness of fit values can be found in Table S2 in the Supporting Material section. Following the first infected individual confirmed in each of the three regions, the numbers of infected individuals in Zhejiang, Guangdong and Xinjiang exhibited a clearly defined first increasing and then decreasing trend. The peak numbers of infected individuals occurred on the 17^th^, 21^nd^, and 22^th^ day after the first case was reported in Zhejiang, Guangdong and Xinjiang, respectively.

Similarly, the transmission, cure and mortality rates in South Korea, Italy and Iran were calculated by the SEIR model, and they are denoted by dot-lines in the top right parts of Figs 3, 4 and 5, respectively. In South Korea, in particular, the transmission rate initially displayed a remarkable bio-peaked pattern and then decreased until a low level was reached. The second period of transmission rate increase begun on February 14^th^, 2020, and it reached its peak (0.278) on February 18^th^. The cure rate reached its peak on February 15^th^, and then decreased rapidly until it reached a low level. Since March 1^st^, 2020, the cure rate started displaying an increasing trend. In Italy, the transmission rate initially increased rapidly and reached its peak (0.262) on February 18^th^, 2020, and then it decreased until it reached a stable level. The cure and mortality rates in Italy exhibited an increasing-fluctuating trend until they reached a stable level. In Iran, the transmission rate reached the highest value (0.142) on February 26^th^, 2020, and it displayed a decreasing trend since then. The cure rate was irregular shown its first peak on February 25^th^, 2020, followed by a second peak on March 6^th^, then it was reduced showing several local (fluctuating) peaks. The mortality rate in Iran reached its first peak on February 21^st^, 2020, then decreased considerably but continued to fluctuate.

Due to the various control and prevention measures taken in South Korea, Italy and Iran to control disease spread, the transmission, cure and mortality rates in these countries exhibited irregular variations. Therefore, these irregularly varying rates were not fitted to any empirical models and were directly inserted in the SEIR model to calculate case numbers. The goodness of fit of the calculated case numbers compared to the empirical case numbers are shown in Table S2 (the *R*^2^ values of the number of infected individuals were around 0.999). The curves (lines) fitted to the empirical numbers of infected, accumulated cure and accumulated death cases can be found at the bottom-right part of Figs 3, 4 and 5, respectively. The disease case numbers curves showed significant differences between South Korea *vs*. Italy and Iran. Until March 11^th^, 2020, the numbers of infected cases in the three countries displayed a constantly increasing trend, although the increment pace of infected cases in South Korea had slowed down by that time; between March 11^th^ and 29^th^, the infected case numbers in South Korea decreased considerably, but in Italy and Iran they were still increasing. The accumulated cured case numbers in South Korea started increasing sharply on March 10^th^, 2020. On the other hand, in Italy and Iran, the accumulated cured case numbers started increasing on March 13^th^ and March 1^st^, respectively, but these increases were very slow. The accumulated death case numbers in South Korea were kept at a low level, were in Italy and Iran, they started increasing on March 13^th^, 2020, and March 10^th^, respectively, and they are still increasing.

Lastly, we simulated the effect of imposing stringent control measures in each country. We selected a date in each country when major disease control measures were implemented. As the timetable in Fig 1 shows, the government of South Korea imposed maximum blockade measures in the Daegu and Gyeongsangbuk-do areas on February 25^th^, 2020. The Council of Ministers in Italy took urgent measures to prevent and control COVID-19 on February 25^th^; and the government of Iran forbade any gatherings on March 8^th^. As noted earlier, we assumed that South Korea, Italy and Iran had taken the equally effective disease control measures as did the Zhejiang, Guangdong and Xinjiang regions, respectively. The simulated transmission rate curves for South Korea, Italy and Iran are denoted by the blue lines at the top right parts of Figs 3, 4 and 5, respectively (the curves in South Korea and Italy start on February 25^th^, and in Iran on March 8^th^, as appropriate). Moreover, the cured rate in the three countries were assumed to be 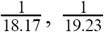, and 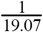, respectively, according to Fig 1 (the number of days between the green and red lines, i.e., 18.17, 19.23, 19.07 days, are the days needed to cure the disease in South Korea, Italy and Iran, respectively, under the simulated conditions); and zero mortality rates were assumed. Using these simulated rates to run the SEIR model, it was found that the number of infected cases in South Korea, Italy and Iran would had reached their highest values on March 9^th^, 10^th^ and 20^th^, respectively, and the corresponding numbers of infected individuals would had been 4480, 2335 and 6969 cases. These simulated infected case numbers in the three countries are much smaller than the actual reported numbers of infected cases observed in South Korea (7212), Italy (8514), and Iran (11466) during the same days. Actually, the differences are striking. For example, the observed maximum infected case number in South Korea on March 11^th^ was 7470, which is also much larger than the maximum simulated infected case number (4480). Moreover, if the effective control measures implemented in Zhejiang were also used in South Korea, the number of days required to reach the peak number of infected cases could have been reduced to two days. Also, the observed numbers of infected cases in Italy on March 29^th^ were 73880, which is more than 30 times larger than the maximum simulated infected case number (2335). The number of the observed infected case number in Italy still has not reached its peak value on March 29^th^. Similarly, the observed infected case number in Iran on March 29^th^ was 23278, which is more than 3 times larger than the maximum simulated infected case number (6969), and it was still increasing on March 29^th^. It is noteworthy that the number of infected cases mentioned above refers to the infected cases still in hospitals, which excludes the number of cured and dead cases. Moreover, we also compared the accumulated number of confirmed cases (the sum of infected, cured and dead case numbers) between the simulated and the actual situations in the three countries up to March 29^th^; the results (observed vs. infected accumulated case numbers) were as follows: 8076 *vs*. 9661 in South Korea, 4199 *vs*. 97689 in Italy, and 15096 *vs*. 38309 in Iran, i.e., a reduction in the accumulated number of infected cases 1585, 93490 and 23213 for South Korea, Italy and Iran, respectively, accounting for 16.41%, 95.70% and 60.59% of the accumulated number of infected cases in these countries.

### 3.4 Comparison of the COVID-19 epidemic control dates in the six regions

Our focus here is to determine when it should be considered that the COVID-19 epidemic was under control (epidemic control date, ECD). For this purpose, we used the transmission, cure and mortality rates to calculate *R*_0_(*t*), and the numbers of infected and exposed cases to calculate *L*_0_(*t*) in each one of the six regions. The epidemic would be considered under control at the time *t*_*ECT*_ (epidemic control time, ECT) after which Eq (3) holds. Two cases were considered: (*a*) the *R*_0_(*t*) and *L*_0_(*t*) curves were plotted and the dates *t*_*ECT*_ were computed based on the actual preventive measures implemented in each region; and (*b*) the *R*_0_(*t*) and *L*_0_(*t*) curves were plotted and the dates *t*_*ECT*_ were simulated in the three countries using the same measures as those implemented in the corresponding Chinese regions. The numerical results are shown in Fig 6.

**Figure 6:**
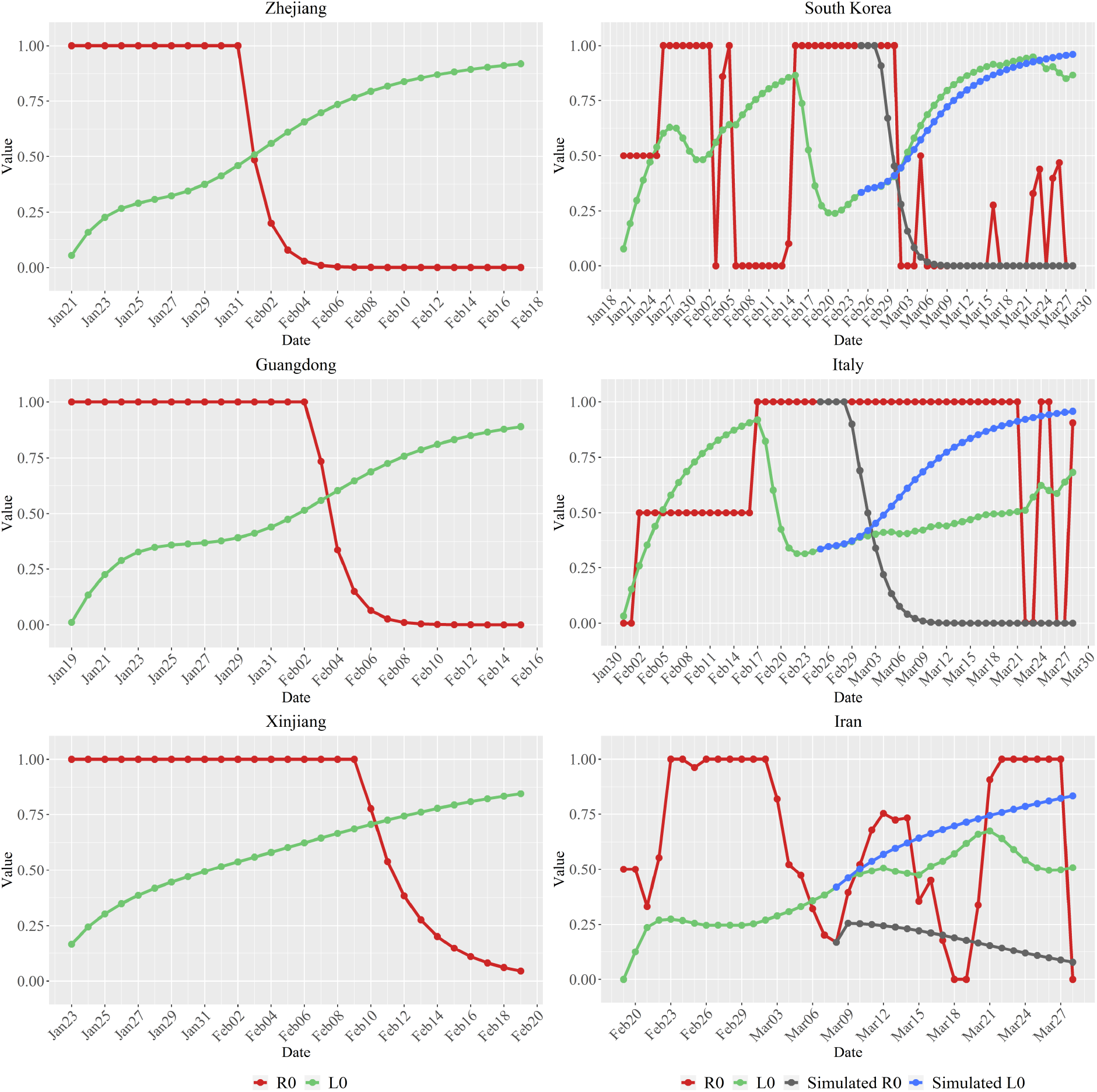
The trends of *R*_0_(*t*) and *L*_0_(*t*) variations in each of the six regions (for better visualization, we set *R*_0_(*t*) = 1 whenever the *R*_0_(*t*) value was computed to be greater than 1).

As can be seen in Fig 6, in the three Chinese regions the inequality of Eq (3) holds on the date *t*_*ECT*_ =February 1^st^, 2020 (Zhejiang), *t*_*ECT*_ =February 4^th^, 2020 (Guangdong) and *t*_*ECT*_ =February 11^th^, 2020 (Xinjiang); i.e., after 9, 12 and 17 days the 1^st^-Level Response to Major Public Health Emergency was declared at Zhejiang, Guangdong and Xinjiang, respectively, and 3, 5 and 3 days before the number of infected cases reached the peak values. This means, that the COVID-19 should have been under control in these regions after theses *t*_*ECT*_ dates (the epidemic reached an “inflection point” at the corresponding dates).

Regarding South Korea, Italy and Iran, both the actual and simulated *R*_0_(*t*) and *L*_0_(*t*) curves are plotted in Fig 6. The actual *R*_0_(*t*) and *L*_0_(*t*) curves are more irregular than in the three Chinese regions. This may happen because the control and prevention measures implemented in these countries were initially rather loose and not as strict as in the Chinese regions. Specifically, under the control and prevention measures actually taken in South Korea the *R*_0_(*t*) and *L*_0_(*t*) curves fluctuate around each other until the inequality *R*0 (*t*) < *L*0 (*t*) is definitely satisfied starting on *tECT* =March 2^rd^, 2020, 9 days before the number of infected cases reached the peak values, which means that the COVID-19 epidemic was effectively under control by that date (the epidemic reached an inflection point). In Italy and Iran the *R*_0_(*t*) and *L*_0_(*t*) curves continued fluctuating around each other since March 20^th^, 2020, and no definite *t ECT* dates could be determined at least until the end of March, 2020. Overall, there is a profound difference between the variation patterns of the *R*_0_(*t*) and *L*_0_(*t*) curves observed in the three Chinese regions and those observed in the three other countries: in the Chinese regions there are well defined *t*_*ECT*_ dates, which is not the case in South Korea, Italy and Iran (i.e., the *R*_0_(*t*) and *L*_0_(*t*) curves fluctuate around each other).

If, on the other hand, it is assumed that the measures in Zhejiang, Guangdong and Xinjiang were implemented in South Korea, Italy and Iran, respectively, the following simulated results are obtained: in South Korea the simulated epidemic control date will be *t ECT* =March 2^rd^, 2020, i.e., the same as the actual date; for Italy, the simulated *t ECT* =March 3^rd^, 2020; and for Iran the simulated *t ECT* =March 8^th^, 2020.

## 4. Discussion

COVID-19 is an infectious disease with largely unknown characteristics but with apparently very considerable societal impacts. Accordingly, everyone needs to make one’s own contribution to its prevention and control. Otherwise, the cost in human and material resources can be very high before the disease can be finally put under control. With the current lack of effective drugs and vaccination, the alternative way to control disease spread is the timely detection of the infection source and cutting off its transmission paths. Chinese authorities proposed a slogan for fighting against the COVID-19: “Quarantine all who need to be quarantined, medically treat all who need to be treated, test all who need to be tested” [7]. In order to explore the behavior of COVID-19 spread under various disease control measures, the current study employed a modified SEIR model to quantitatively assess the variations of the temporal trajectory of COVID-19 spread in three regions of China as well as in three COVID-19 affected countries.

### 4.1 The COVID spread pattern in the three regions of China

The measures taken to control the COVID-19 spread in China during the early transmission period (including limiting social contacts and outside activities as much as possible, quarantining the infected individuals and their close contacts as soon as possible) were shown to effectively prevent the disease from spreading to larger groups of people by lowering the transmission rate. With such control measures, the transmission rates reached their peaks within one or two days, and were reduced to zero within 11, 11 and 18 days after a 1^st^-Level Response to Major Public Health Emergency was declared in Zhejiang, Guangdong, and Xinjiang, respectively. As more knowledge was gained about clinical treatments, the cure rate increased and the mortality rate decreased during the first 28 days of the COVID-19 outbreak.

It was also found that a normal distribution function adequately represented the variation of the transmission rate in the Zhejiang, Guangdong and Xinjiang regions of China under the above control measures. But the transmission rate at Xinjiang exhibited considerable fluctuations around the normal distribution model, and it took more time (18 days) for the transmission rate to drop to zero than it did in Zhejiang (11 days) and Guangdong (11 days). This may be associated with the advanced socio- economic development in Zhejiang and Guangdong. Also, the governance capability in Xinjiang is different from that in Zhejiang or in Guangdong, which led to varied enforcement degrees of disease prevention and control measures in these regions. Some clues can be found in Fig 1, i.e., after Xinjiang launched the 1^st^-Level Response to Major Public Health Emergency, it seems likely that the most stringent prevention and control measures were only taken initially in the largest cities. Then, on January 28^th^, 2020, Xinjiang strengthened the improved treatment of patients at the county level. Furthermore, on February 1^st^ Xinjiang intensified its implementation of the prevention and control in rural areas. All these actions may be reflected in the fluctuations of the transmission curve (Fig 5).

### 4.2 The spread patterns in the three countries

The disease transmission rates in South Korea, Italy and Iran during the study period experienced much higher peak values (0.278, 0.262 and 0.142, respectively) than in Zhejiang, Guangdong and Xinjiang (0.096, 0.075 and 0.053, respectively), suggesting that some special events took place that caused these higher peaks in the three countries. Specifically, the Daegu Church gathering in Daegu city, and the infections in Daenam hospital (Gyeongsangbuk-do province) were found to be two major events that produced large infected populations, which comprised 82% of the accumulated infected cases on March 29^th^, 2020. In the same day, it was reported that 84.1% of the accumulated infected cases were related to group outbreaks [19]. On February 22^nd^, an infected individual was confirmed in Lombardia (Italy) who had not traveled abroad. This was a key indicator that COVID-19 had spread domestically. Given that the disease incubation period can be several days long, it is expected that the number of confirmed patients will increase only after the virus is transmitted among people. Moreover, the medical staff was unaware of the highly infectious nature of COVID-19; therefore, a large number of medical staff got sick in Lombardia. Up to March 29^th^, the number of infected cases in Lombardia accounted for 42.23% of the infected cases nationwide. In Iran, although the transmission rate decreased significantly during the period from February 26^th^ to March 8^th^, 2020, unfortunately the rate increased again after March 8^th^, indicating that the disease was not yet under control. The apparent high cure rate in Iran could be linked to the relative loose hospital rules that allowed the early release of patients. Due to inadequate medical resources in these hospitals, the patients were sent to isolation places for further monitoring after leaving the hospital.

Moreover, the simulated tests performed in the present study concerning disease spread in South Korea, Italy and Iran were based on two modeling conditions: (*a*) the disease transmission law implemented in the three Chinese regions was also used for the three countries under consideration, and (*b*) similar measures as in the three Chinese regions were assumed to have been taken to control the disease spread at these three countries. Based on these conditional disease simulations, it was found that the COVID-19 spread in South Korea, Italy and Iran could be controlled significantly, resulting to a much smaller infected cases and a shorter time to reach the maximum number of infected individuals (bottom right parts of Figs 3-5). If effective measures could be implemented at the early stage of the COVID-19 outbreak, the number of infected cases would be controlled and kept at relative low levels, which will reduce the burden on the medical system; in return, the population health risk, the mortality rate, and the public health costs could be considerably reduced at both the nationwide and the local levels. For example, the mortality rate and the accumulated infected case numbers in the three regions of China are much smaller than those of the three countries considered. Specifically, the mortality rates during the first 28 days of the COVID-19 outbreak in Zhejiang, Guangdong and Xinjiang were 0%, 0.15%, 1.32%, respectively. On the other hand, the mortality rates in South Korea, Italy and Iran were 1.64%, 11.03% and 6.89%, respectively, during the period since the first reported case date to March 29^th^. In addition, the difference between the simulated and the observed accumulated infected case numbers on March 29^th^ in South Korea was smaller than those in Italy and Iran, suggesting that the control measures taken in South Korea had better effects on prevention and control than in the other two countries, and that the prevention and control measures in Italy and Iran could had been considerably improved. It is reported that infection control measures such as national lockdowns in many European countries can lead to a reduction of the number COVID-19 related deaths between 21000 and 120000 at the end of March [31].

### 4.3 Disease control measures in China

Below, the disease control measures taken in China are briefly reviewed with references to local authorities and communities, the medical system, the industry, and the prevailing societal and individual perspectives [32]. By implementing these measures, the resulting drop in travel was as high as 90% during the period from January 23^rd^ to February 18^th^, 2020 compared to the same period (lunar calendar) in 2019 (additional information can be found in the Supporting Materials section).

Naturally, a country’s medical society is at the forefront of the war against an epidemic. In the COVID-19 case, the main measures taken by the medical society are aimed at the timely and accurate diagnosis of the disease, and the best possible treatment or curing for the infected individuals. The specific measures are listed in Table S3. Given the rather long COVID-19’s incubation period and the fact that even asymptomatic infections can be substantial, the local governments needed to act fast and do their best to control the interactions of local people. Three key points should be highlighted, in this respect: avoid importing infected individuals in a region, control the local disease spread, and prevent exporting infected individuals to other areas. Table S4 lists the main measures implemented by local governments during the COVID-19 pandemic. Citizen communities are the basic contributing units in a country’s overall effort to prevent a disease from spreading further. Therefore, among basic functions of a local community in China were to assist in identifying infected patients, restricting community access, and providing the basic needs of daily life to its members. The specific measures taken by local Chinese communities are listed in Table S5. The contribution of a country’s industrial sector can offer considerable support in the effort to control an epidemic. Some of the actions taken by the country’s industry are listed in Table S6. Lastly, the best way for an individual to contribute is to be self-isolated and pay more attention to personal hygiene. Some basic guidelines are as follows: wash hands frequently, wear a facemask when going outside, and stay at home for a 14-day self-isolation period.

## 5. Summary and Future Research

The modified SEIR model was shown to provide an adequate framework to represent the patterns of COVID-19 transmission in Zhejiang, Guangdong, and Xinjiang of China, as well as in the countries of Italy, South Korea and Iran. The proposed SEIR model accounts for the interplay of medical, physical and social processes. Not only it can make useful predictions, but it can also offer guidance about what disease spread parameters should be controlled and in what way in order to prevent undesirable epidemic results. The early implementation of the control and prevention measures can lead to a lower number of infected cases and a lighter burden on the medical system, which, in return, can lead to a larger cure rate and a smaller mortality rate.

The temporal moving window scheme helps characterize the dynamics of the COVID-19 transmission rates in these regions during the disease outbreak, which can also benefit the public health managements. The transmission rates associated with the control measures in the three Chinese regions under study (including the immediate quarantine of the infected patients and their close contacts, and the considerable restrictions on social contacts) followed a modified normal distribution function. Thus, in terms of the experience gained in China concerning the prevention and control of disease spread, self-isolation was a key measure of controlling the spread and the transmission. Furthermore, our simulation results showed that if the SEIR epidemic model parameters for South Korea, Italy and Iran were selected so that they corresponded to the disease control measures implemented in China on February 25^th^, February 25^th^ and March 8^th^, respectively, then considerably better results would had been obtained in the three countries (up to March 29^th^, the simulated reduction in the accumulated infected case numbers would had been 1585, 93490 and 23213 for South Korea, Italy and Iran, respectively, accounting for 16.41%, 95.70% and 60.59% of the accumulated number of infected cases), including the flattening and shrinking of the infected case curves, which would also had peaked at an earlier time than the observed curves in these countries. This means that the disease could have been controlled faster and more efficiently in these countries than it actually happened.

Beyond being used to quantitatively describe the current transmission conditions in the six study regions worldwide, the proposed approach has the potential to monitor disease transmission rates and predict disease case numbers in future situations. Health authorities could assimilate this valuable information into their disease prevention and control decision-making process. Future work would also focus on employing the dynamic transmission rate to forecast any trends in the numbers of COVID-19 cases, or to model patterns in future epidemics. Furthermore, spatiotemporal disease characteristics (including the composite space-time disease dependencies and spread patterns, and their association with climatic factors [33-37]) could be explored by considering county-level or even individual-level disease data.

## Data Availability

Data used in this manuscript was collected from the Health Commissions of China and other countries.

## Acknowledgement

We would like to thank Mr. Jimi He (The Chinese University of Hong Kong, Shenzhen) for the assistance with the R software coding. We are very grateful to Drs. Carlos M. Duarte and Susana Agusti (King Abdullah University of Science and Technology) for their encouragement. This work was supported in part by the National Science Foundation of China (41671399) and the Department of Science and Technology, Zhejiang Province (2016C04004).

## Supporting Material

**Figure S1:**
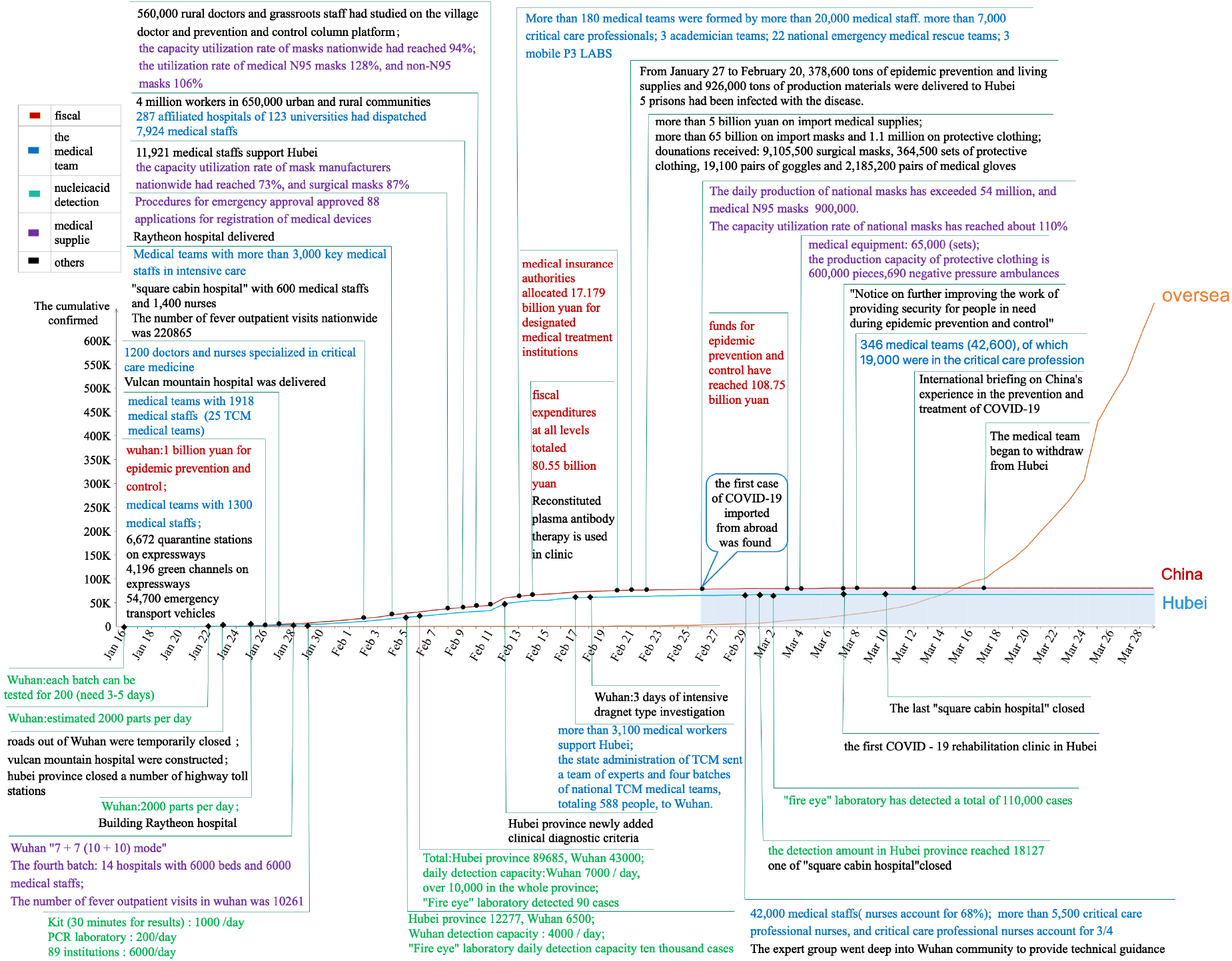
Number of confirmed COVID-19 cases in Hubei Province, mainland China and outside China with the corresponding combined governmental and societal efforts. The data were obtained from [1-3].

**Figure S2:**
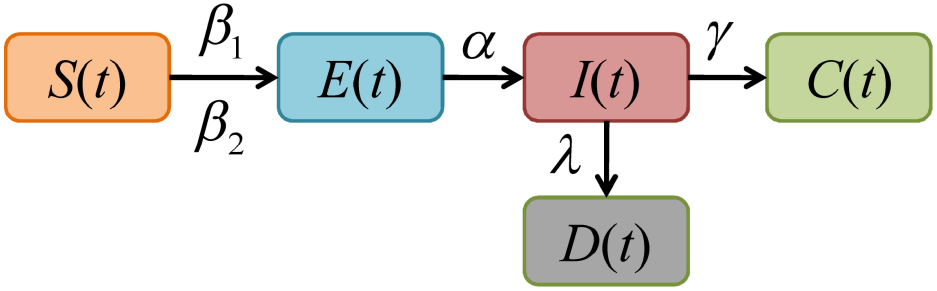
An outline of the SEIR model

### Text S1. Description of the six study areas

Zhejiang province is located in eastern China (27°06′N-31°11′N and 118°01′E-123°10′E), and has a territory of approximately 0.10 million km^2^ and a population of 54 million [4]. Residents between 15- 59 years old and older than 60 years account, respectively for about 72.90% and 13.89% of the population. Zhejiang has a typical subtropical monsoon climate with an annual mean temperature ranging from 15 to 18 °C and an annual mean precipitation range from 980 to 2000 *mm* [5]. The GDP of Zhejiang during 2018 was about 5.62 trillion CNY, which ranked fourth in China.

Guangdong province is located in southern China (20°13′N-25°31′N and 109°39′E-117°19′E), with an area of about 0.18 million km^2^ and population of about 104 million. About 76.36% of the Guangdong population consists of people 15-64 years old, and 6.75% of the population is older than 65 years. Guangdong’s climate belongs to the East Asian monsoon region. More specifically, the northern, middle and southern parts of Guangdong are middle subtropical, south subtropical and tropical areas, respectively. The annual mean precipitation ranged from 1300 to 2500 *mm* with an average value of 1777 *mm* [6]. The GDP of Guangdong in 2018 was 9.72 trillion CNY.

With an area of 1.66 million *km*^2^, Xinjiang Uygur Autonomous Region is located on the northwestern border of China (73°20′-96°25′E and 34°15′-49°10′) with a temperate continental climate. The annual mean precipitation is about 147 *mm*. 22 million people live in Xinjiang, including 55 nationalities, among which about 58% are Muslims. 73.02% of the population ranges in age from 15 to 54 years old, and 6.19% is older than 65 years. The GDP of Xinjiang in 2018 was about 1.22 trillion CNY.

The Republic of Korea (South Korea) is located in the southern part of the Korean peninsula, with an area of about 0.10 million *km*^2^ [7]. A single nationality consisting of 52 million people live in South Korea, among whom 50% are religious. The climate in South Korea is temperate zone monsoon. The annual mean temperature ranges from 13-14 °C and the annual mean precipitation ranges from 1300-1500 *mm*. In 2018, South Korea’s GDP was 1.54 trillion dollar and the per capita national income was 31 thousand dollar.

The Republic of Italy is located in the southern part of Europe with the area of about 0.30 million *km*^2^ [7]. The climate in most parts of Italy belongs to the subtropical Mediterranean region. More specifically, the mean temperature ranges from 2-10°C during January and 23-26°C during July. In 2018, about 60 million people lived in Italy, among whom 22.6% were older than 65 years, and 13.4% were younger than 14 years. The GDP of Italy (1.75 trillion euro) ranked fourth in Europe and eighth in the world. Italy’s per capita national income is 29.0 thousand dollar.

The Islamic Republic of Iran (Iran) is located in the southwestern part of Asia, with an area of 0.16 million *km*^2^ [7]. There are about 81 million people in Iran and most of them live in Tehran, Isfahan, Fars, Khorasan Razavi and East Azerbaijan. Persians make up 66% of the population, Azerbaijanis 25%, Kurds 5%, and the rest are Arabians, Turkmens and others. Islam is the state religion in Iran. The climate in Iran belongs to the continental region, characterized by cold winters and hot summers. Most areas are dry with little rain. In Teheran, the capital of Iran, the hottest month of the year is July, with mean minimum and maximum temperature of 22 °C and 37 °C, respectively; and the coldest is January, with mean minimum and maximum temperatures of 3 °C and 7 °C, respectively. In 2018, Iran’s GDP was 430 billion dollar.

### Text S2. The modified normal distribution function and the quadratic function

In order to explore the variation pattern of the disease transmission, cured and mortality rates, the empirical values of the three rates need to be fitted to a theoretical model. Specifically, the modified normal distribution model was used ([8]),

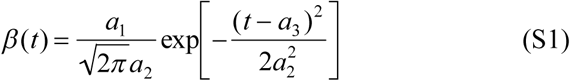

where *a*_1_, *a*_2_ and *a*_3_ denote the scale, shape and location parameters, respectively. Additionally, the cured and the mortality rates were fitted to quadratic equations as

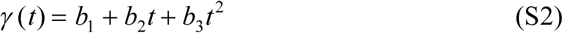

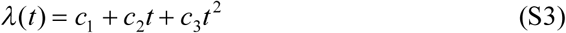

where *b*_1_ and *c*_1_ are constants, and the *b*_2_, *b*_3_, *c*_2_ and *c*_3_ are parameters.

Using the “nlinfit.m” function in the MATLAB package, the parameters of the modified normal distribution equation and quadratic models were calculated as shown in Table S1.

**Table S1.** The values and the corresponding 95% confidence interval of the fitted models

| Model | Parameter | Zhejiang | Guangdong | Xinjiang |
| --- | --- | --- | --- | --- |
| Modified normal distribution model | $a_1$ | 8.13E-01<br>(7.15E-01, 9.11E-01) | 8.24E-01<br>(7.68E-01, 8.80E-01) | 1.64E+00<br>(-6.63E-01, 3.94E+00) |
| | $a_2$ | 3.38E+00<br>(2.88E+00, 3.89E+00) | 4.12E+00<br>(3.78E+00, 4.46E+00) | 1.29E+01<br>(2.39E+00, 2.34E+01) |
| | $a_3$ | 4.30E+00<br>(3.86E+00, 4.74E+00) | 7.20E+00<br>(6.89E+00, 7.51E+00) | -2.84E+00<br>(-2.30E+01, 1.74E+01) |
| Quadratic model | $b_1$ | -4.89E-03<br>(-1.38E-02, 4.02E-03) | 6.96E-03<br>(-4.64E-04, 1.34E-02) | 7.47E-03<br>(1.79E-04, 1.48E-02) |
| | $b_2$ | 7.74E-04<br>(-6.43E-04, 2.19E-03) | -2.23E-03<br>(-3.27E-03, -1.20E-03) | -2.67E-03<br>(-3.83E-03, -1.51E-03) |
| | $b_3$ | 7.08E-05<br>(2.34E-05, 1.18E-04) | 1.56E-04<br>(1.21E-04, 1.90E-04) | 1.62E-04<br>(1.23E-04, 2.00E-04) |
| Quadratic model | $c_1$ | -2.04E-04<br>(-1.12E-03, 7.06E-04) | 5.75E-26<br>(3.03E-26, 8.48E-26) | 1.93E-03<br>(-1.53E-04, 4.02E-03) |
| | $c_2$ | 2.06E-05<br>(-1.24E-04, 1.65E-04) | -1.05E-26<br>(-1.48E-26, -6.17E-26) | -5.33E-04<br>(-8.64E-04, -2.01E-04) |
| | $c_3$ | 1.44E-06<br>(-3.41E-06, 6.28E-06) | 3.45E-28<br>(2.00E-28, 4.90E-28) | 2.42E-05<br>(1.32E-05, 3.53E-05) |

### Text S3. The passenger transportation situation

Chinese Spring Festival, or New Year, is the world’s largest annual migration. The high peak travel period during the Spring Festival of 2020 was 40 days, from January 10^th^ to February 18^th^. Due to the emergency measures described earlier, the estimated travel drop could be as high as 90% [9].

- Rail passengers: A total of about 210 million passengers were transported during the period from January 10^th^ to February 18^th^ of 2020. For the first 15 days (January 10^th^ to 24^th^), a total of about 169 million passengers were transported, an increase of 17.2% (24.63 million) passengers compared to the same period in 2019. For the remaining 25 days (January 25^th^ to February 18^th^) since the implementation of the national major public health emergency, a total of 42.48 million passengers were transported, a reduction of 220 million passengers and a drop of 83.9% compared to the same period of 2019. Assuming that passenger transportation would follow the same increasing pattern between the first 15 days and the remaining 25 days during the entire travel period, we estimated that the passenger travel drop due to COVID-19 could be more than 90%.
- Air passengers: According to the statistics obtained from the Beijing Capital Airport, a total of about 5.7643 million passengers have transited this airport during the period from January 10^th^ to February 18^th^ in 2020, a reduction of about 5.7 million passengers and a drop of 50% compared to the same period in 2019. For the period between January 10^th^ and 23^rd^, the daily average of air passengers was about 0.28 million. Immediately following the implementation of the national major public health emergency, air passengers experienced a sharp drop and maintained at very low numbers. Thus, we estimated a drop of about 90% in passenger transportation by air. This could well represent a national level estimation too.
- Bus and automobile passengers: We have not found statistics on passenger travel by the bus and over the highway system. However, the majority of passenger buses between provinces and between cities were suspended after the implementation of the national major public health emergency. On February 24, 2020, among the 31 provinces, autonomous regions and metropolitan areas of the Mainland, a total of 27 provinces resumed inter-provincial bus transportation. Then, we estimated that, generally, the drop in passenger travel by the bus system could be even more than that by rail or air.

**Table S2:**
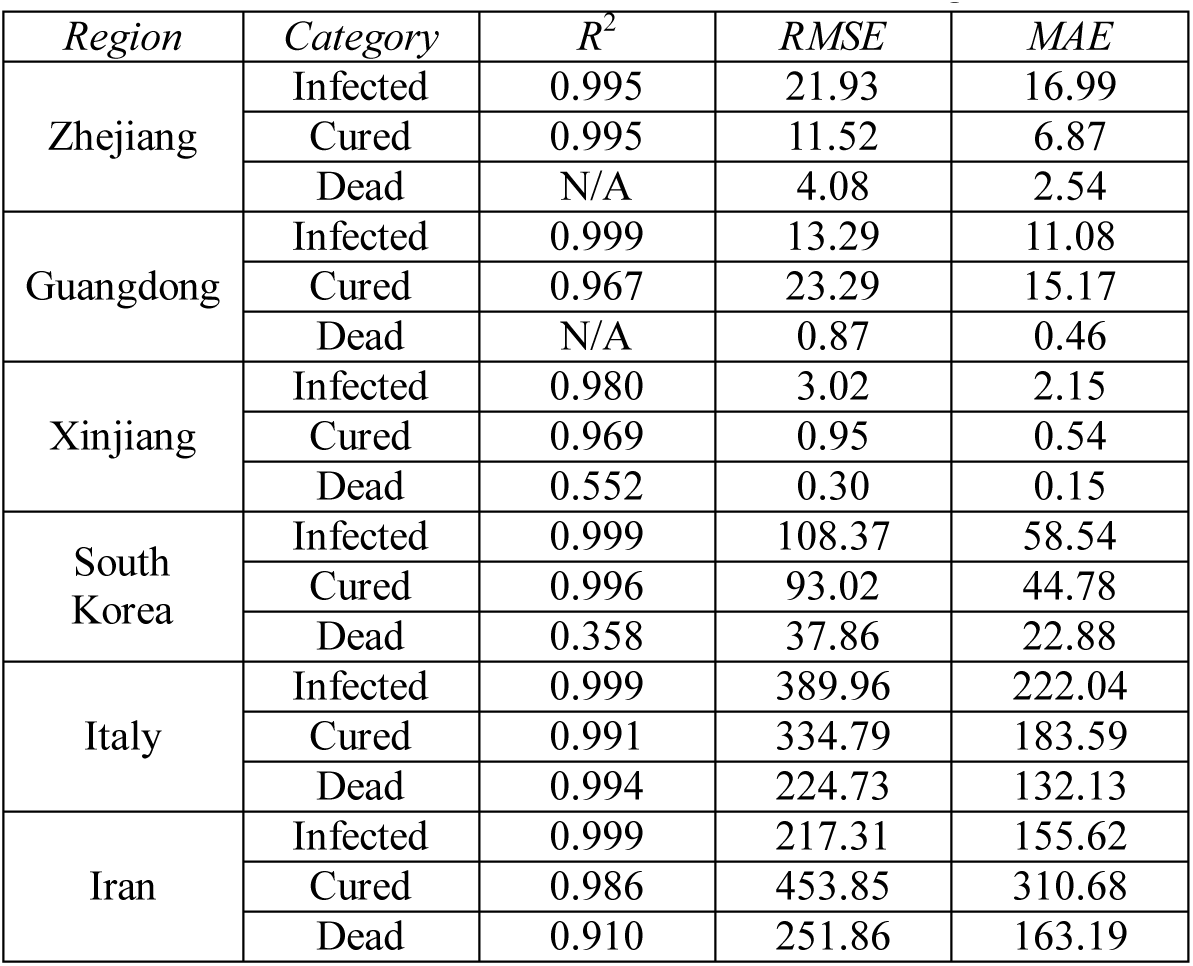
Goodness of fit of SEIR modeling.

**Table S3:**
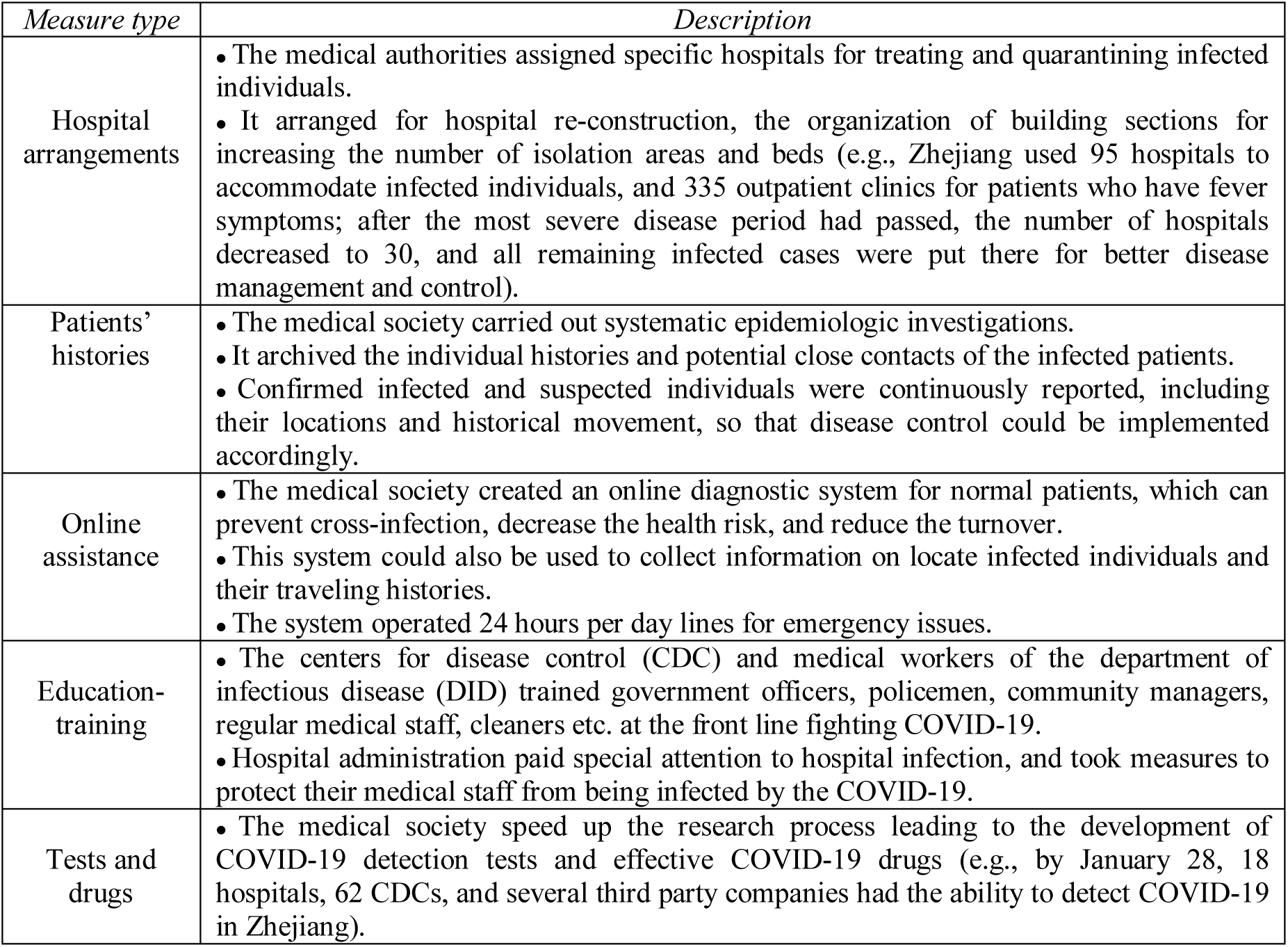
Medical society’s measures (China) [1-3,10].

**Table S4:** Local government measures (China) [1-3,10].

| *Measure type* | *Description* |
| --- | --- |
| Immediate restrictions | • Public places (schools, parks, restaurants, malls, cinemas, libraries etc.) were closed.<br>• Public and family gatherings (*in-situ* meetings, cultural and sports activities, family feasts etc.) were cancelled.<br>• Public transportation (buses, ships, planes) between provinces, cities and counties were suspended.<br>• Places closely associated with people's everyday lives were strictly monitored (e.g., food markets were disinfected twice a day and the body temperatures of all people entering them were measured). |
| Check-points | • Check-points were setup at all city and county entrances (e.g., at various points of highway exits) and at all train station and airport exits.<br>• Police and medical staff monitored the body temperatures of all people passing through check-points, and their origins were registered (e.g., people who lived in epidemic-affected areas or whose temperature was above normal were sent to hospitals for further testing, diagnosing or quarantining).<br>• During the most severe epidemic period, only people with local identification cards and certified local workers could pass the check-points at the county and city entrances. |
| Resources | • Special funds were timely allocated to control local disease spread, prepare medical facilities, purchase protective equipment, and further respond to disease transmissions (e.g., by January 27<sup>th</sup>, Zhejiang authorities had provided more than 1.2 billion yuan to support disease control actions).<br>• Sufficient staff was timely transferred to support local communities and medical systems in order to prevent disease spread (e.g., by January 26<sup>th</sup>, i.e. 3 days after Zhejiang announced a 1<sup>st</sup>-Level Response to Major Public Health Emergency, more than 330 thousand officers contributed to disease control together with community managers).<br>• If the number of infected people was too large and there were not enough hospital beds available, public places (e.g., gymnasiums, theatres and music-halls) were restructured to serve as shelter hospitals.<br>• Additional specific guidelines were setup to separate household waste from medical facilities waste and other relevant places hosting patients. |
| Health-codes | • A 3-color health code was used to manage the local flow of people by identifying the condition of people entering a local county or city (Zhejiang was the first province to use this code).<br>• People applied for health code in specific cities based on their location, current health condition and the places they had been during the past two weeks.<br>• A red health code meant that an individual was infected, or suspected, or had closely contacted an infected (or suspected) individual, or had possibly contacted an infected (or suspected) individual, or came from a severely epidemic-affected region. In all cases, people were quarantined for 14 days, reporting their health condition every day. After that, an individual's code would turn into green.<br>• A yellow health code meant that one came from a severe disease region or had potentially contacted someone from that region. In this case, one needed to be quarantined for 7 days, having to report one's health condition during each day. After that, an individual's code would turn to green.<br>• A green health code meant that the individual had very low risk of getting the disease and was free to travel.<br>• Schools and companies also used this code to monitoring the health condition of students, teachers and staff.<br>• Local governments, schools and companies may have more restricted codes for allowing their staff to return to these places and maintain their basic functions. |
| Asymptotic treatment | • A sufficient number of places were arranged for asymptomatic people who had, though, been in close contact with infected individuals, and the necessary health staff was assigned to these places.<br>• Hotels, public houses and school dormitories were requisitioned for these people reside in.<br>• Health-care teams (each consisting of 1 officer, 1 doctor and 2 nurses) were put together to take care of 50-people groups. The doctor and nurses monitored people's temperature and overall condition 2-3 times a day, and those whose body temperature exceeded a threshold or exhibited other disease symptoms were sent to hospitals for further treatment. |
| Scientific indexes | • A scientific index was used to assess each county's or city's health risk, and subsequent control strategies were implemented at each county or city. |
|  | • Zhejiang used 5-color maps to assess each county's health risk by considering the accumulated number of infected individual, ratio of the number of local patients over total number of patients, clustered cases, the number of continuous days that no new infected individuals were detected (e.g., Zhejiang announced that the disease control focus switched from preventing the import of infected individuals to avoiding local disease spread, and its efforts switched from disease control to both disease control and ensuring economic development at the same time). |
| Legal punishment | • People whose actions threatened public health were legally punished (e.g., people who hide their health condition, violated local disease control measures, or concealed that they were coming from or traveling to areas at a severe disease state). |

**Table S5:** Local communities’ measures (China) [1-3,10].

| *Measure type* | *Description* |
| --- | --- |
| Community health duties | • Among the community managers' duties were to make sure that everyone who was in a public place was wearing a face-mask.<br>• They were also instructed to continuously assess their community's status, including the daily health condition and travel history of every community member.<br>• Managers classified their community members into four groups: of low concern were members who completed the isolation period without any symptoms, of current focus were member still within the isolation period, newly arrived (i.e., today) members from other places should be body temperature tested and closely monitored, and early prevention measures should be taken for the member who arrived recently (i.e., yesterday or earlier) from other places.<br>• During the isolation period, the managers visited their community members and measured their body temperature 3-times a day. If a member's temperature was above the threshold or some typical symptoms of illness occurred, the member was sent to the nearest designated hospital, and the closely contacted members should be home-quarantined for further monitoring (e.g., in a Hangzhou city community, a group of 339 managers visited 12025 families in 334 buildings, collected detailed information about them, and 122 disease-related clues were detected for the isolated families). |
| Community check-points | • Check-points were setup to restrict community access 24 hours per day. Only community members could pass through the check-points after body temperature testing.<br>• During the most severe disease period, one member per family could leave the community every 2 days to purchase food. |
| Community assistance | • Managers assured satisfactory living conditions for their community members.<br>• During the isolation period, the community assisted all its members, especially senior ones, with their daily food purchases.<br>• A 24 hours a day emergency response system was established at each community. |
| Disinfection | • Community managers arranged for the disinfection of public areas, physical exercise facilities, garbage centers, elevators, apartment entrances etc. |
| Community information-Instruction | • Managers kept their community members informed about the current COVID-19 status and the necessary individual safeguard procedures.<br>• Managers used moving broadcasting or unmanned aerial vehicles for community information purposes.<br>• Managers instructed their community members to avoid unnecessarily gathering, and they shut down community facilities (e.g., swimming pools, libraries etc.) |

**Table S6:**
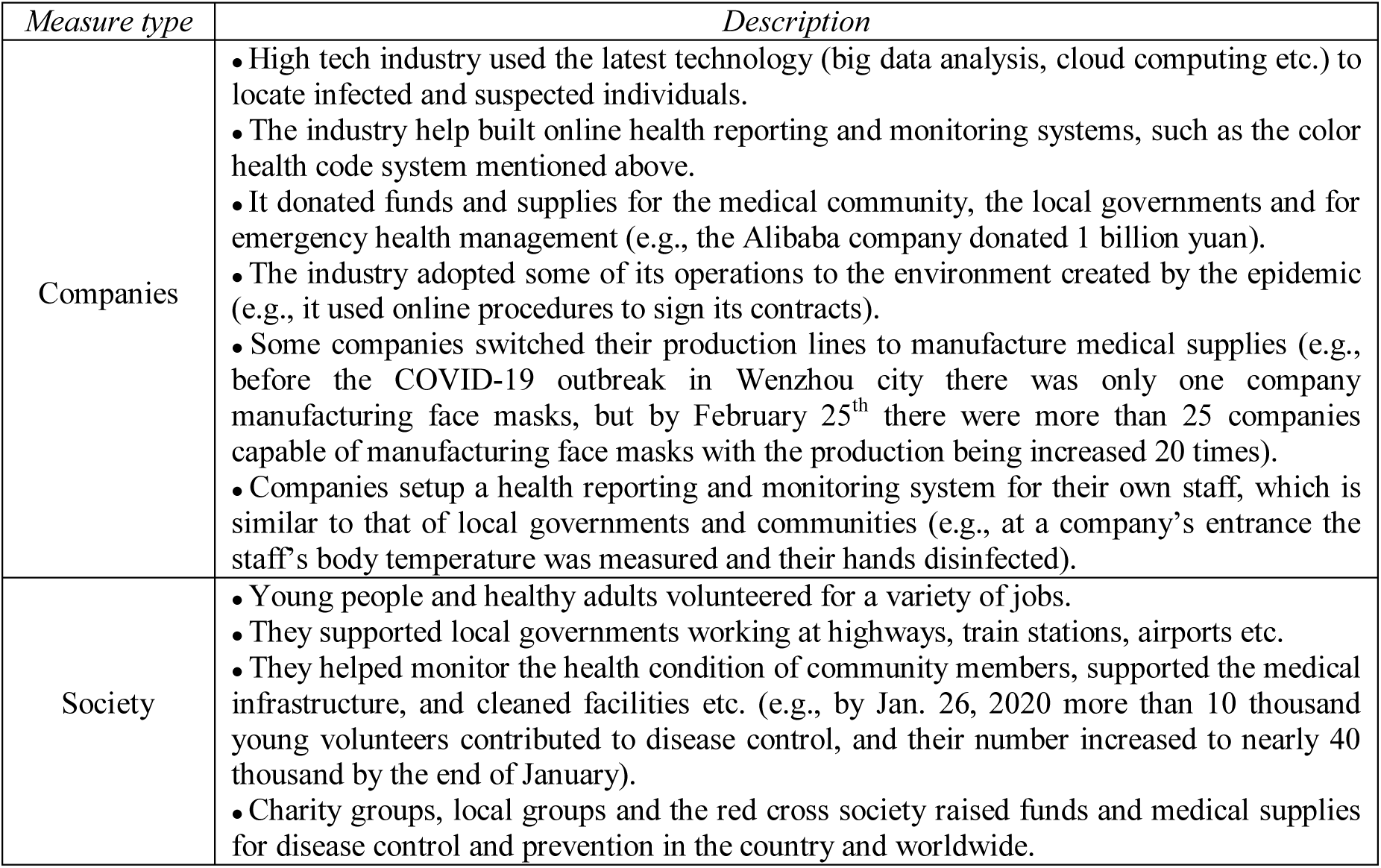
Industrial sector’s measures (China) [1-3,10].

| *Measure type* | *Description* |
| --- | --- |
| Companies | • High tech industry used the latest technology (big data analysis, cloud computing etc.) to locate infected and suspected individuals.<br>• The industry help built online health reporting and monitoring systems, such as the color health code system mentioned above.<br>• It donated funds and supplies for the medical community, the local governments and for emergency health management (e.g., the Alibaba company donated 1 billion yuan).<br>• The industry adopted some of its operations to the environment created by the epidemic (e.g., it used online procedures to sign its contracts).<br>• Some companies switched their production lines to manufacture medical supplies (e.g., before the COVID-19 outbreak in Wenzhou city there was only one company manufacturing face masks, but by February 25<sup>th</sup> there were more than 25 companies capable of manufacturing face masks with the production being increased 20 times).<br>• Companies setup a health reporting and monitoring system for their own staff, which is similar to that of local governments and communities (e.g., at a company's entrance the staff's body temperature was measured and their hands disinfected). |
| Society | • Young people and healthy adults volunteered for a variety of jobs.<br>• They supported local governments working at highways, train stations, airports etc.<br>• They helped monitor the health condition of community members, supported the medical infrastructure, and cleaned facilities etc. (e.g., by Jan. 26, 2020 more than 10 thousand young volunteers contributed to disease control, and their number increased to nearly 40 thousand by the end of January).<br>• Charity groups, local groups and the red cross society raised funds and medical supplies for disease control and prevention in the country and worldwide. |

